# Statin Initiation and early stroke recurrence in the POINT Trial Population

**DOI:** 10.1101/2024.05.07.24307030

**Authors:** Elham Azizi, Shyam Prabhakaran, James R. Brorson

## Abstract

**Background and Purpose:** Benefits of statin therapy in reducing the long-term risk of ischemic stroke are well-established, but the immediate effects of statin therapy on early stroke recurrence after an initial ischemic event are less clear.

**Methods:** In this secondary analysis of POINT data, we evaluated the effects of statins on early stroke recurrence (within 7 days) and recurrence over 90 days. We also examined the effect of early statin initiation in the subgroup of subjects not on statins prior to the index event using logistic and proportional hazards models.

**Results:** In the POINT trial, 175 of 267 (65.5%) of ischemic stroke recurrences were early (within 7 days). Baseline statin treatment at the time of study entry was marginally associated with a decrease in the odds of early ischemic stroke recurrence in adjusted logistical regression analysis (OR 0.70, 95% CI 0.50-0.99, p=0.04), but with a non-significant effect in adjusted Cox proportional hazard analysis (HR=0.72, 95% CI 0.52–1.01, p=0.05). In the subset of subjects not taking statin medications at baseline, initiation of statin treatment had no significant effect on the rate of early stroke recurrence (adjusted HR 0.80, 95% CI 0.54–1.20, p=0.22).

**Conclusion:** In the POINT trial population, prior treatment with statin was weakly associated with decreased risk of early recurrence of stroke. However initiating statin treatment had no detectable effect in reducing risk of early stroke recurrence. The POINT trial provides no evidence for an immediate protective effect of statins against stroke recurrence.

## Introduction

After an initial minor stroke or transient ischemic attack (TIA), the risk of recurrence of stroke is highest within first few days. Trials of various antiplatelet regimens for acute secondary prevention initiated within 12 to 24 hours following the index event, such as the Clopidogrel in High-Risk Patients with Non-disabling Cerebrovascular Events (CHANCE)^1^, Acute Stroke or Transient Ischaemic Attack Treated with Aspirin or Ticagrelor and Patient Outcomes (SOCRATES)^2^, Platelet-Oriented Inhibition in New TIA and Minor Ischemic Stroke Trial (POINT)^3^, and Acute Stroke or Transient Ischemic Attack Treated with Ticagrelor and ASA for Prevention of Stroke and Death (THALES)^4^, have been consistent in showing that the early recurrences within the first week exceed those in following months.

The long-term benefits of statin therapy in reducing risk of ischemic stroke are well-established. For example, in primary stroke prevention in patients with history of cardiac disease the Heart Protection Study showed that simvastatin treatment was associated with a substantial reduction in the rate of ischemic stroke and TIA^5^. In secondary stroke prevention, the SPARCL trial (Stroke Prevention by Aggressive Reduction in Cholesterol Levels) confirmed that atorvastatin 80 mg/d reduced recurrent stroke rates in patients with history of recent TIA or stroke but without cardiac risk factors, across all baseline ischemic stroke subtypes^6^.

The short-term effects of statins on early stroke recurrence after an initial stroke or TIA are less clear. Retrospective analysis has suggested that prior treatment with statins improves stroke survival, especially when statins are resumed within 2 days of the stroke, and that statin withdrawal at the time of stroke is associated with increased mortality^7^. A prospective observational study showed that statin pretreatment in patients with large artery atherosclerotic strokes was associated with lower 1-month stroke recurrence and mortality^8^. However small trials of early versus late initiation of atorvastatin 80 mg were unable to demonstrate significant differences in the growth of infarction volume or in the rate of ischemic stroke recurrence^9,10^.

The POINT trial^3^, which examined the effects of dual antiplatelet therapy with clopidogrel and aspirin compared to monotherapy with aspirin in minor acute ischemic stroke and high-risk TIA, provides unique data regarding stroke recurrence from a very early time point following minor stroke or TIA, with subjects randomized with 12 hours of the initial event. Statin treatment status of subjects was recorded at baseline and at 7 day and 90 day follow up visits. We sought to investigate whether this dataset could provide evidence for the effects of statin treatment on early stroke recurrence, and in particular, whether immediate initiation of statin treatment in those not treated with statins prior to the event could prevent the high rate of early recurrences following initial minor stroke or TIA.

## Methods

We used the STROBE cohort checklist when writing our report ^11^.

### Clinical datasets

The POINT trial dataset was provided by the National Institute of Neurological Disease and Stroke. The shared datasets included 4,881 subjects from the POINT trial. The present study was submitted to the University of Chicago Institutional Review Board and was determined to be exempt from further review.

The POINT trial compared dual antiplatelet therapy with clopidogrel and aspirin to monotherapy with aspirin alone, tracking outcomes including a primary composite endpoint (IS, MI, and vascular death) as well as safety outcomes. Subjects in the control group were treated with aspirin alone, while those in the active treatment groups received loading doses of aspirin and clopidogrel followed by aspirin 81 mg daily and clopidogrel 75 mg daily, for 90 days.

### Statistical methods

Survival tables were prepared from the shared datafiles and statistical analyses were performed using SAS Studio version 9.4. Intention-to-treat populations, definitions of outcome events, designation of event times, and data censoring methods were defined according to the trial protocol. The present analysis utilized treatment assignments by the intention-to-treat principle, and examined ischemic stroke recurrence as the outcome event, testing for effects of baseline statin treatment status, and for effects of early statin initiation. For the latter, a variable coding for statin initiation by the 7 day visit was created and assigned the value of the day 7 statin treatment status for subjects without a censoring or failure event before 7 days, or, if this value was missing, the value of the baseline statin treatment status. For subjects with censoring or failure events before day 7, the outcome event statin treatment status was used, or if this was missing, the baseline statin status was used. Statin treatment status was missing in 2 subjects at baseline, in 137 subjects at 7 days, and in 389 subjects at the 90-day visit.

Modifying effects of several variables were studied, including the presence of cervical carotid disease, cohort (those with minor stroke versus TIA as initial events), treatment assignment, systolic blood pressure, serum glucose, age, and race. Known carotid disease was dichotomized as present for stenosis > 50% on either side, or as absent, with missing values for 681 subjects imputed a value of 0 (no known disease). Race was also dichotomized, assigning a value of ‘1’ for categories of Black/African American or American Indian/Alaska Native (categories previously found in univariate analysis to be significantly associated with risk of early recurrence) or ‘0’ for other categories, including White, Asian, and ‘Other’, and leaving 133 subjects with missing values. Missing or unknown values for other variables included hypertension history 21, diabetes mellitus 9, serum glucose 3, and systolic blood pressure 4.

Binary logistical regression was performed, using complete case analysis, excluding subjects with missing values for any variable. 153 subjects censored before 7 days (most on the day of randomization) were excluded from this logistic regression analysis. Previous multivariate logistic regression analysis, that had included these early censored subjects, and had imputed values of ‘0’ for missing values of relevant variables, had shown significant associations (p<0.01) of recurrent ischemic stroke with presence of known carotid disease, patient cohort, treatment assignment to placebo, baseline systolic blood pressure, baseline serum glucose, age, prior statin use, and dichotomized race^12^. The present logistic regression analysis adjusted for these variables.

For Cox proportional hazard analysis, the effect of baseline statin treatment on time to ischemic stroke recurrence was modeled both over the entire 90-day trial period and with a focus on the initial 7 days. The analysis again adjusted for presence of known cervical carotid disease, cohort, treatment assignment, systolic blood pressure, serum glucose, age, and race. Subjects were then divided between those on and those not on statin treatment at baseline, and effects of statin initiation before the day 7 visit on early stroke recurrence was examined, with adjustment for the same variables.

### Subgroup analysis

Of particular interest in acute management of acute stroke patients is whether early initiation of statin in those not treated with statin at baseline might protect against early recurrence. As POINT included a large group of subjects not on statin treatment at baseline who were started on statin treatment between the baseline and 7-day visits, we tested whether statin initiation following the baseline visit might affect risk of stroke recurrence. The dataset was divided between those on statin at baseline and those not on statin at baseline, and adjusted Cox proportional hazard analyses were performed on the group of patients not on statin treatment at baseline, in which there were 118 ischemic stroke events in 2899 subjects. Attribution of statin initiation was based on recorded statin treatment at the 7-day visit or, for events occurring before 7 days, at the outcome event visit, or, if these values were missing, on the baseline statin treatment status.

## Results

In POINT, most (267 of 281) composite outcome events were ischemic stroke, and these recurrences of ischemic stroke occurred early, with 175 occurring within 7 days of trial entry, and only 92 occurring over the remainder of the 90-day trial. In the POINT trial population, only a minority of 1890 of 4881 (39%) of subjects were on statin treatment at baseline, while at the 7-day visit, 3713 of 4723 (79%) were treated with statins.

Binary logistic regression analysis on effects of baseline statin treatment status on any recurrence of ischemic stroke over 90 days was performed. In unadjusted regression analysis, the effect of baseline statin treatment was not significantly associated with risk of stroke recurrence over 90 days (OR 0.83, 95% CI 0.64-1.08, p=0.16), but in adjusted analysis baseline statin treatment was associated with reduced risk (OR 0.72, 95% CI 0.55–0.95, p=0.02). Similarly, focusing on early ischemic stroke recurrence within 7 days of study entry, in unadjusted analysis the effect of baseline statin treatment again was not significantly associated with risk of stroke recurrence (OR 0.76, 95% CI 0.55–1.05, p=0.09), but in adjusted logistic regression there was a modest independent association of baseline statin treatment with reduced odds of early recurrence within 7 days (OR 0.70, 95% CI 0.50-0.99, p=0.04). Known carotid disease, cohort (minor stroke rather than TIA), systolic blood pressure, treatment assignment, serum glucose, and race were also independently associated with early recurrence (Table).

Cox proportional hazard analysis was applied to examine the effect of statin treatment on the time to recurrence of ischemic stroke. Over the full 90-day study period, baseline statin treatment exhibited a non-significant trend towards lowering the risk of stroke recurrence in unadjusted analysis (HR 0.83, 95% CI 0.65–1.07, p=0.15), and in adjusted analysis, a modest protective effect (adjusted HR=0.73, 95% CI 0.56–0.96, p=0.02).

Analysis focusing on early recurrence in the initial 7-day period showed a similar point estimate of a modest protective effect of baseline statin treatment that did not reach statistical significance in either unadjusted analysis (HR 0.76, 95% CI 0.56–1.04, p=0.09) or in adjusted analysis (adjusted HR=0.72, 95% CI 0.52–1.01, p=0.05; Figure 2).

**Figure 1.**
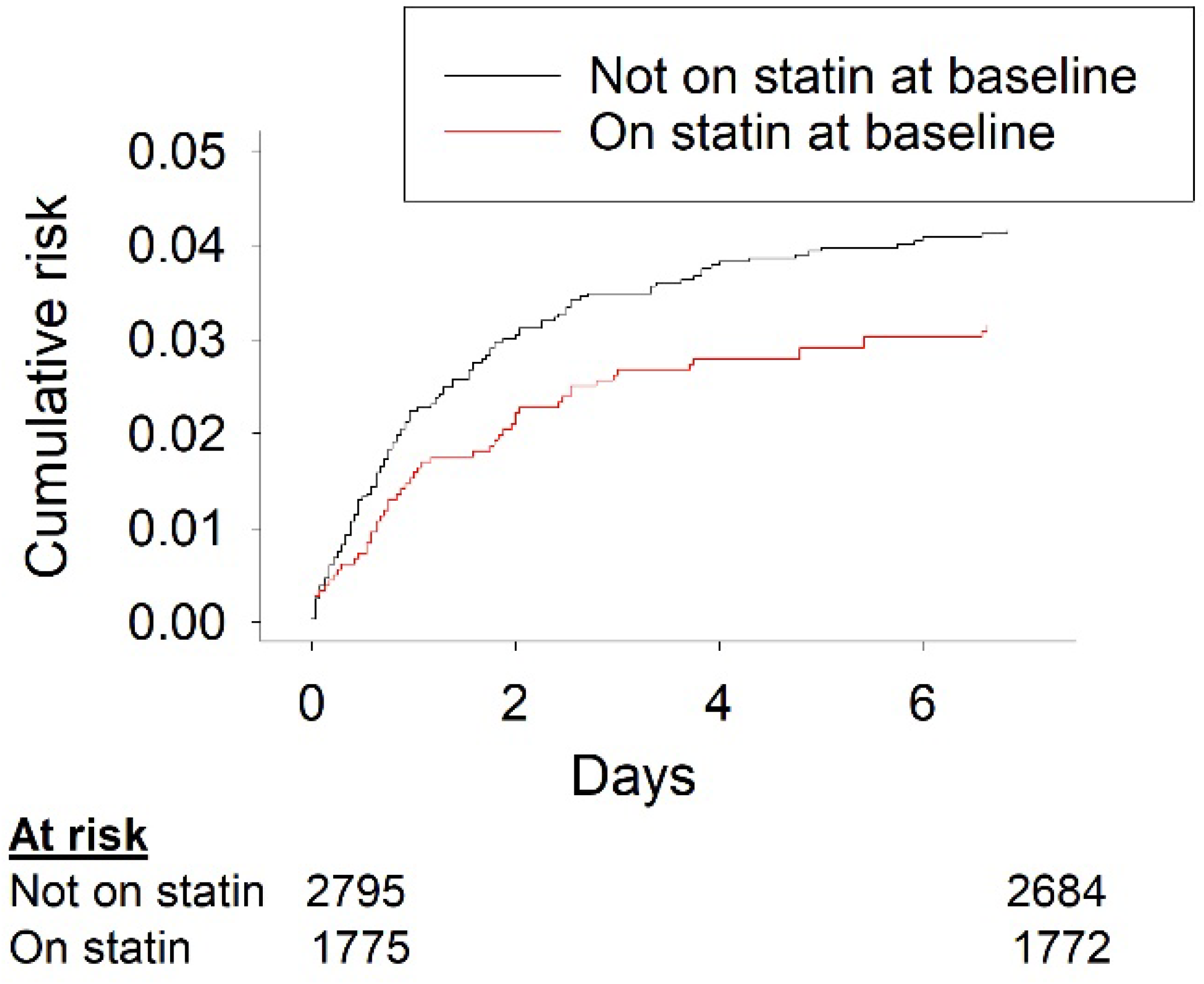

**Figure 2.**
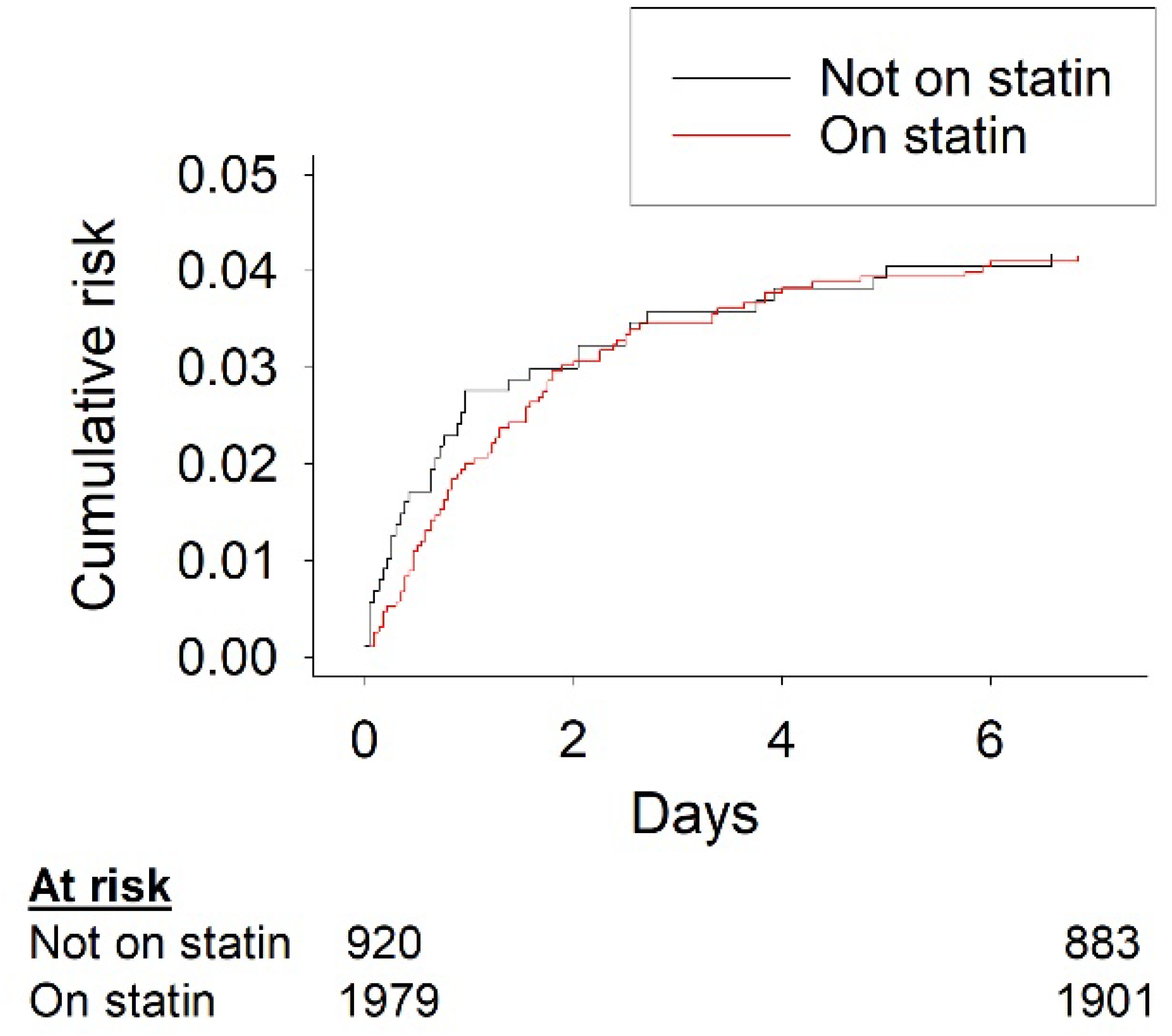

In 920 subjects not initiated on statin treatment, 38 events occurred while 80 events occurred in 1979 subjects initiated on statin treatment. In both unadjusted and adjusted analyses, there was no evidence for any effect of early initiation of statin treatment on the risk of early recurrent within 7 days (unadjusted HR 0.98, 95% CI 0.66–1.43, p=0.9; adjusted HR 0.80, 95% CI 0.54– 1.20, p=0.22; Figure 3).

## Discussion

In the POINT trial population of patients with minor ischemic stroke or TIA, prior treatment with statin was weakly associated with decreased risk of ischemic stroke recurrence, but initiating statin treatment in those not previously on statins had no significant effect on the risk of stroke recurrence within the first 7 days. This analysis provides no support for any immediate protective effect of statin treatment on the risk of early stroke recurrence. Any mechanism that accounts for the long-term protective effect of statins appears to be one that develops over more than 7 days after statin initiation.

As expected, based on known ischemic stroke preventive effects of statin treatment^12^, baseline statin treatment was found to be associated with reduced odds of both early ischemic stroke recurrence and recurrence over the extended 90-day period. The estimated protective effects of baseline statin were similar over the 7-day and 90-day periods, perhaps because most of the subjects on statin treatment at baseline continued to be treated with statin throughout their time in the trial. While not recorded in the dataset, serum cholesterol levels were likely to be lower in subjects on statins at baseline. Effects of lower baseline lipid levels on recurrence of stroke in patients with symptomatic intracranial atherosclerotic disease have previously been observed over the 6-8 weeks following the initial event^13^.

In a systematic review, Amarenco et al. found that the stroke risk reduction provided by statins correlated with the reduction in LDL-C they produced, with a risk reduction of over 15% per 10% reduction in LDL-C^13^. The lipid-lowering effects of oral administration of statins such as atorvastatin develop over several days’ time, are incomplete at 7 days but are near-maximal at 14 days^15^. Serum lipid levels are thought to modulate cholesterol deposition in vascular plaque, with intimal lipid accumulation, macrophage infiltration, and inflammatory responses contributing to plaque destabilization that leads to thromboembolism and stroke^16^. If statins’ protective effect is occurring primarily through lipid-lowering mechanisms, the time course of these subsequent mechanistic steps, in addition to the delay in achievement of lipid-lowering, could explain the lack of an immediate protective effect of starting a statin.

In addition to the multiple studies describing associations with lipid-lowering of statin effects on stroke prevention and progression of large artery stenosis, some studies suggest the importance of other ‘pleiotropic’ statin effects. One small study compared doses of simvastatin and of ezetimibe producing similar LDL-C reductions in heart failure patients, and demonstrated evidence of improved endothelial function by several independent assays with simvastatin, and not with ezetimibe, suggesting that simvastatin improved endothelial function independently of its effect on LDL levels^17^. The effects of simvastatin in dyslipidemic subjects in reducing endothelial dysfunction have been associated with reduced activity of the Rho-associated coiled-coil containing protein kinase (ROCK) pathway^18^.

An attempt to investigate the effect of early intensive statin treatment in acute coronary syndrome patients using simvastatin 40-80 mg versus placebo for the first 4 months failed to show a significant effect on the primary composite clinical outcome endpoint, but suggested an antilJinflammatory effect, in that at 4 months patients in the high-intensity simvastatin group showed reduced C-reactive protein levels, with a trend toward fewer cardiovascular death events^18^. The Pravastatin or Atorvastatin Evaluation and Infection Therapy-Thrombolysis in Myocardial Infarction (PROVE IT-TIMI)^20^ study also has been proposed as supporting the pleiotropic effects of atorvastatin 80 mg, including inflammation modulation, improved endothelial function, and anticoagulation, as some of the reason for early benefits seen within the first 30 days after acute coronary syndrome^21^. These pleiotropic effects are appealing as a mechanism by which plaque inflammation and instability might be suppressed by statins following an initial stroke, potentially in a rapid fashion, preventing early stroke recurrence.

Unfortunately, no direct support of this concept is found in the present analysis of the POINT trial data.

There are significant limitations to this study pertaining to the limits to the data reported in the POINT trial. The timing of statin initiation following stroke or TIA, or the specific drug or dosage used, were not recorded. Although recognized cardioembolic strokes were excluded, the specific stroke mechanism was not defined in each case. Missing values for statin treatment status at the time of outcome events were frequent, requiring the assumption that subjects with early events remained at the status of statin use at baseline. Most importantly, the 95% confidence interval is wide for the hazard ratio for effect of statin initiation on early stroke recurrence, including both the possibilities of no effect and of a substantial reduction in event rates with statin initiation. To more confidently determine whether early statin initiation may be effective in reducing early stroke recurrence, a larger prospective study of this intervention would be required.

## Conclusion

In this secondary analysis of the data of the POINT trial, while the history of prior statin use at baseline is weakly associated with a decreased odds ratio of early recurrence of ischemic stroke within 7 days, there is no evidence for an immediate protective effect of introduction of statin treatment during the 7-day period in those not previously on statins. Further studies are needed to confirm these results, and to explore the influence of timing and route of initiation of lipid-lowering therapy on stroke recurrence following minor stroke or TIA.

## Data Availability

All data produced in the present work are contained in the manuscript

## Acknowledgements

The authors would like to thank the National Institutes of Neurological Disorders and Stroke for provision of the POINT trial dataset.

---

**Table.**
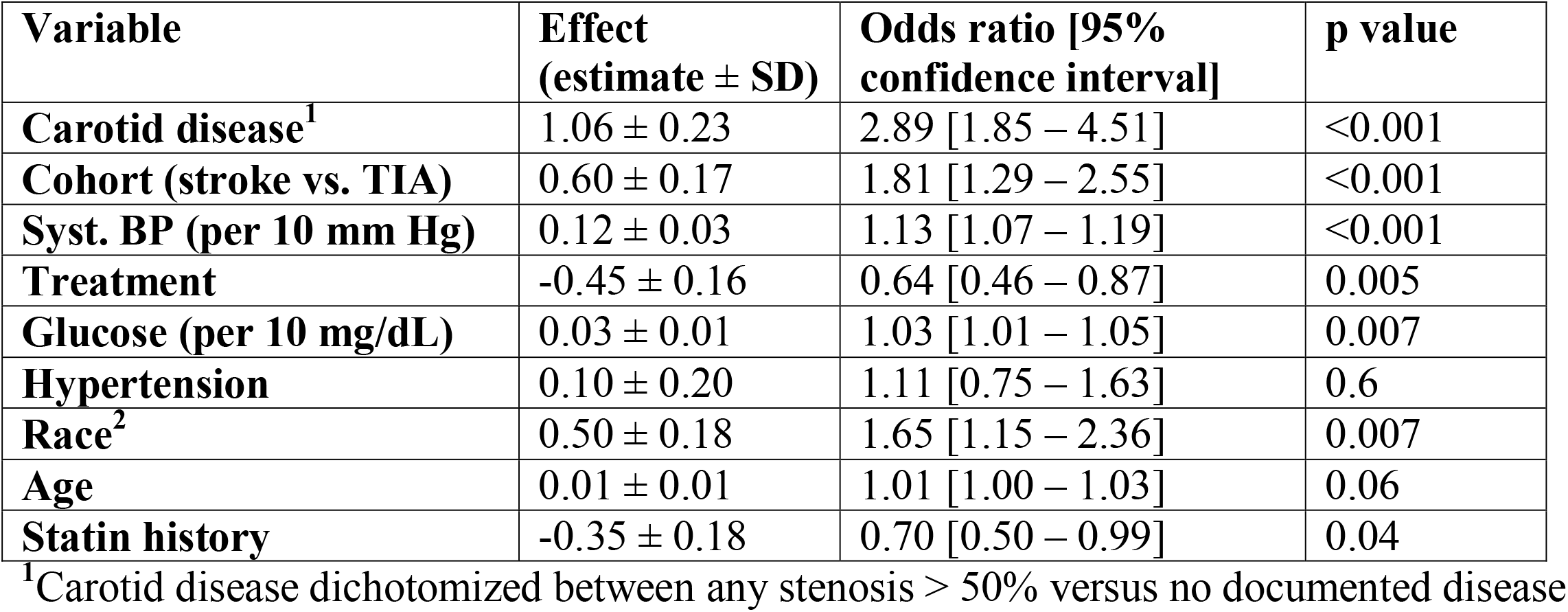
Effect of baseline statin use on recurrence of ischemic stroke within 7 days: Results of adjusted multivariate logistical regression analysis.

**Table.**
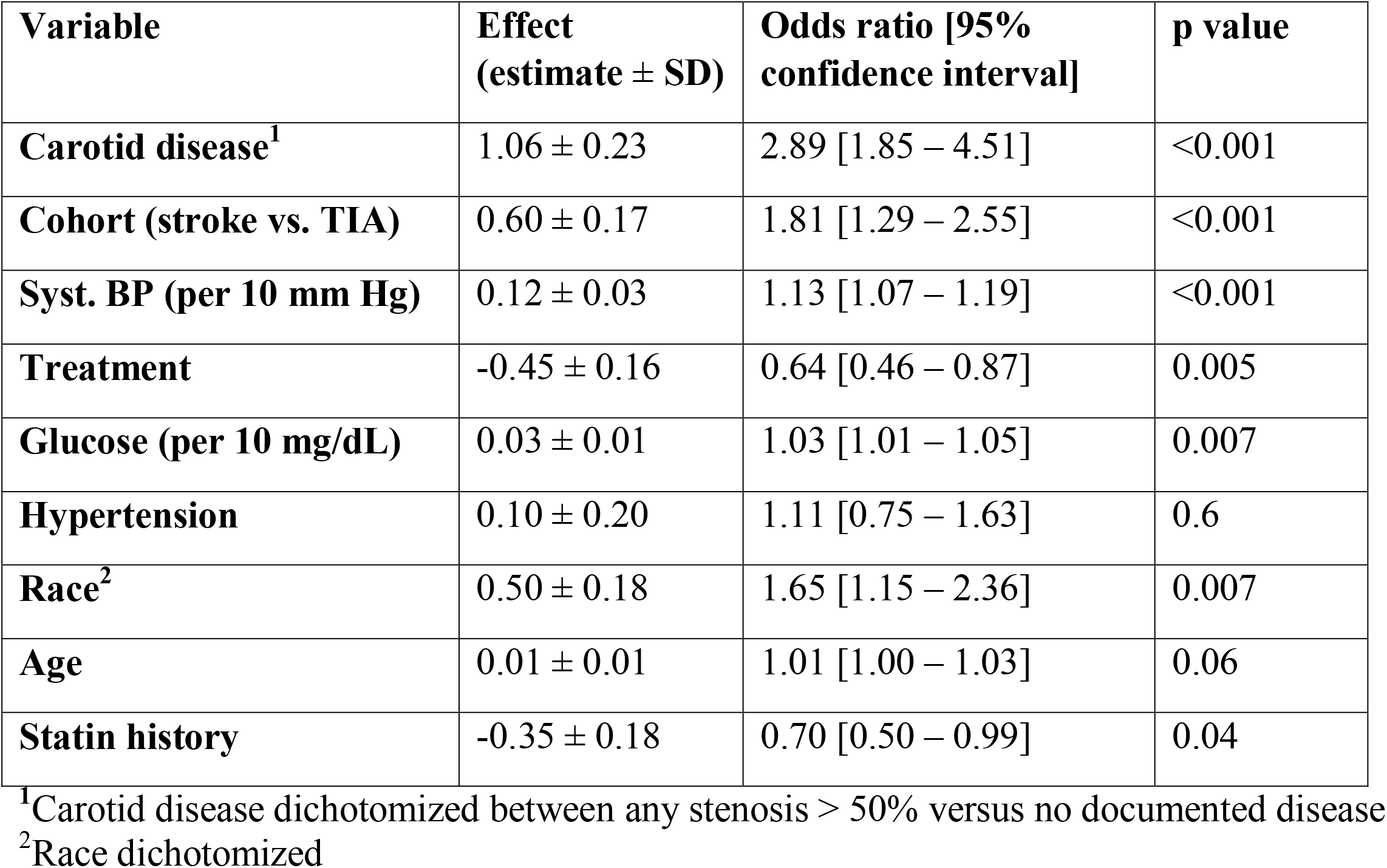
Effect of baseline statin use on recurrence of ischemic stroke within 7 days: Results of adjusted multivariate logistical regression analysis.

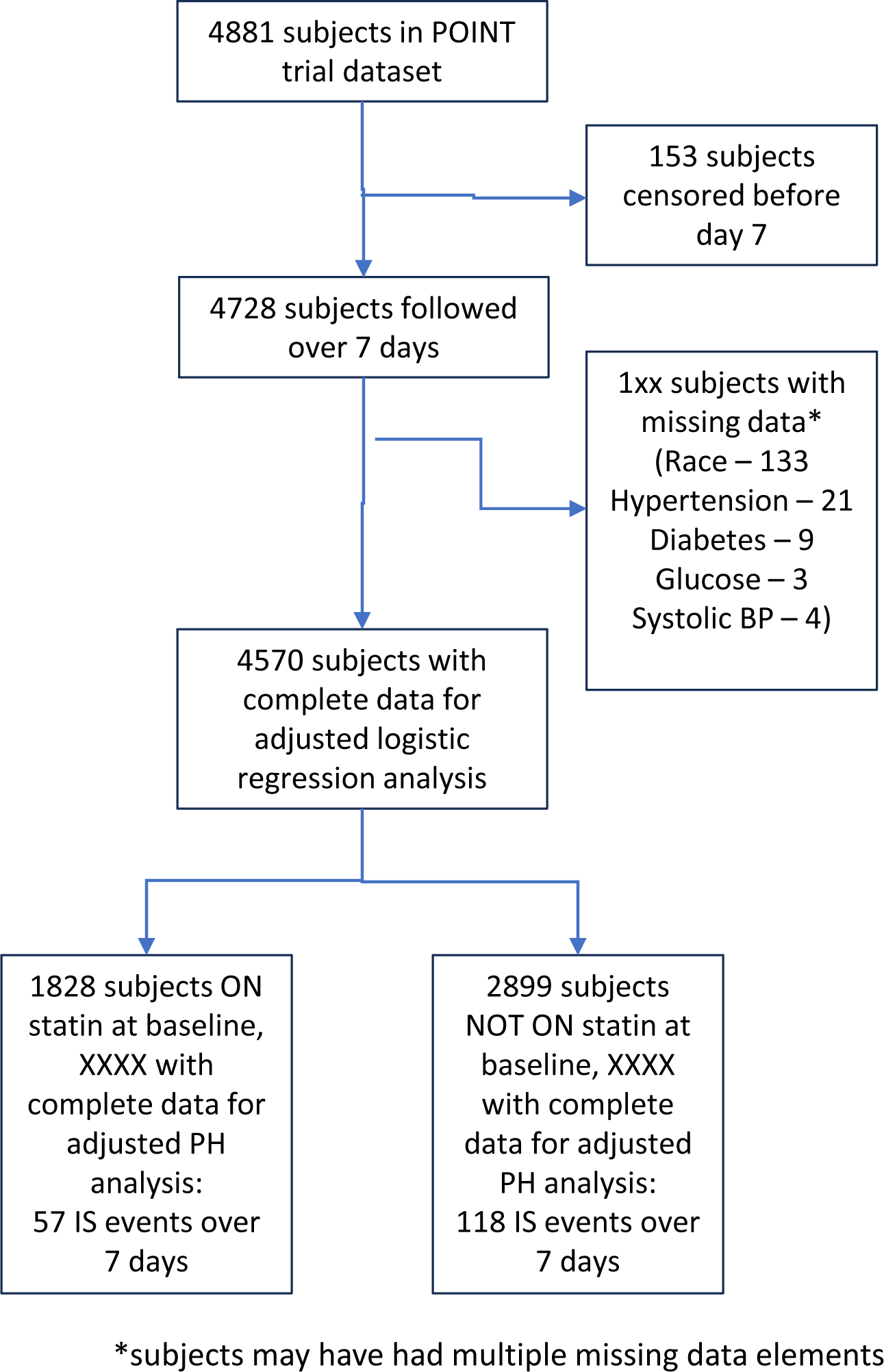

